# Safety and cardiovascular effects of multiple-dose administration of aripiprazole and olanzapine in a randomised clinical trial

**DOI:** 10.1101/2020.08.03.20167502

**Authors:** Dora Koller, Susana Almenara, Gina Mejía, Miriam Saiz-Rodríguez, Pablo Zubiaur, Manuel Román, Dolores Ochoa, Aneta Wojnicz, Samuel Martín, Daniel Romero-Palacián, Marcos Navares-Gómez, Francisco Abad-Santos

## Abstract

**Objective:** To assess adverse events and safety of aripiprazole and olanzapine treatment.

**Methods:** Twenty-four healthy volunteers receiving 5 daily oral doses of 10 mg aripiprazole and 5 mg olanzapine in a crossover clinical trial were genotyped for 46 polymorphisms in 14 genes by qPCR. Drug plasma concentrations were measured by HPLC-MS/MS. Blood pressure and 12-lead ECG were measured in supine position. Adverse events were also recorded.

**Results:** Aripiprazole decreased diastolic blood pressure on the first day and decreased QTc on the third and fifth day. Olanzapine had a systolic and diastolic blood pressure, heart rate and QTc lowering effect on the first day. Polymorphisms in *ADRA2A, COMT, DRD3* and *HTR2A* genes were significantly associated to these changes. The most frequent adverse drug reactions to aripiprazole were somnolence, headache, insomnia, dizziness, restlessness, palpitations, akathisia and nausea while were somnolence, dizziness, asthenia, constipation, dry mouth, headache and nausea to olanzapine. Additionally, *HTR2A, HTR2C, DRD2, DRD3, OPRM1, UGT1A1* and *CYP1A2* polymorphisms had a role in the development of adverse drug reactions.

**Conclusions:** Olanzapine induced more cardiovascular changes; however, more adverse drug reactions were registered to aripiprazole. In addition, some polymorphisms may explain the difference in the incidence of these effects among subjects.

## Introduction

Aripiprazole (ARI) and olanzapine (OLA) are atypical antipsychotics commonly prescribed for patients with schizophrenia or schizoaffective disorders (Robert R. Conley and Kelly, 2005). OLA has a higher serotonin (5-HT)2A/dopamine D2 receptor binding ratio compared to typical antipsychotics. Additionally, it has antagonistic activity at dopamine D3 and D4, 5-HT3 and 5-HT6, histamine H1, α1-adrenergic and muscarinic M1-5 receptors (Bhana et al., 2001; Bymaster et al., 1996). On the contrary, ARI is a partial agonist at dopamine D2, D3, D4 and serotonin 5-HT1A, 5-HT2C and α1-adrenergic receptors and it is a 5-HT2A and 5-HT7 antagonist (Shapiro et al., 2003).

ARI is predominantly eliminated through hepatic metabolism by cytochrome P450 (CYP) isozymes 3A4 and 2D6. Its main active metabolite, dehydro-aripiprazole (DARI) has similar affinity for the dopamine D2 receptor and represents approximately 40% of ARI exposure in plasma (Kim et al., 2008). OLA is mainly metabolized by direct glucuronidation via the UDP-glucuronosyltransferase (UGT) enzyme family, by oxidation via CYP1A2 and secondarily by CYP2D6 and CYP3A4 (Callaghan et al., 1999).

Several typical and atypical antipsychotics increase the risk of heart rate-corrected QT (QTc) prolongation and, as a consequence, *Torsades de Pointes* and sudden cardiac death (Ray et al., 2009). In previous studies with schizophrenic patients, mean QTc interval was decreased with ARI and QTc prolongation risk was lower with ARI and OLA compared to other antipsychotics (Czekalla et al., 2001; Polcwiartek et al., 2015; Ray et al., 2009; Vieweg, 2003). However, although ARI usually does no produce QTc prolongation, some studies reported the contrary in patients and healthy volunteers (Belmonte et al., 2016; Hategan and Bourgeois, 2014; Suzuki et al., 2011). Additionally, patients with impaired CYP2D6 enzyme activity (i.e. poor metabolizers, PMs) may be at a greater risk of QTc prolongation (Dorado et al., 2007).

Neuroleptic Malignant Syndrome, although it is rare, may occur with the administration of all antipsychotics. One of its symptoms is fluctuation in blood pressure (BP) (Sarkar and Gupta, 2017). Therefore, it is of emphasized importance to monitor BP during antipsychotic treatment. ARI and OLA may produce hypotension in both patients and healthy volunteers (Belmonte et al., 2016; Jana et al., 2015; Wang, 2013). Additionally, ARI can induce hypertension (Yasui-Furukori and Akira Fujii, 2013).

Both ARI and OLA can cause an increase in heart rate (HR) (Belmonte et al., 2016; Tajiri et al., 2018). However, when changing the therapy from OLA to ARI, a significant decrease was detected in HR (Tajiri et al., 2018). Therefore, OLA had a stronger HR increasing effect compared to ARI and it was dose-dependent (Tajiri et al., 2018).

The most common adverse drug reactions (ADRs) to ARI in schizophrenic patients are akathisia, extrapyramidal symptoms, somnolence and tremor (Ribeiro et al., 2018). On the contrary, the most common ADRs to OLA in schizophrenic patients are constipation, weight gain, dizziness, personality disorder, akathisia, postural hypotension, sedation, headache, increased appetite, fatigue, dry mouth and abdominal pain (R. R. Conley and Meltzer, 2000).

The aim of the current study was to evaluate the safety of 5 days treatment with ARI and OLA and their effects on electrocardiogram (ECG) and BP. Moreover, we aimed to correlate these factors with sex, plasma drug concentrations and genetic polymorphisms.

## Materials and Methods

### Study Population and Design

Twenty-four healthy volunteers (12 males and 12 females) were enrolled in a multiple oral dose, open-label, randomized, crossover, two-periods, two-sequences, single-centre, comparative, phase I clinical trial performed between June 2018-April 2019. Five doses of 10 mg/day ARI tablets or 5 mg/day film-coated OLA tablets were administered to each volunteer during 5 consecutive days. Block randomization was used to assign a treatment to each volunteer on the first day. The drug was administered at 09:00 h each day under fasting conditions. After a 28 days washout period, each volunteer received the opposite drug. The random allocation sequence, the recruitment of participants and their assignment to interventions were performed by investigators of the Clinical Trials Unit.

The clinical trial was performed at the Clinical Trials Unit of our hospital. The protocol was approved by its Research Ethics Committee authorized by the Spanish Drugs Agency and under the guidelines of Good Clinical Practice and the Declaration of Helsinki (EUDRA-CT: 2018-000744-26). All subjects were adequately informed about the study and, if agreeing to participate, signed an informed consent form before inclusion. The inclusion and exclusion criteria were reported in our previous publication (Koller, Saiz-Rodríguez, et al., 2020).

### Pharmacokinetic Analysis

Twenty-two blood samples were collected in EDTA K2 tubes for pharmacokinetic assessments during each treatment period of the study. Samples were centrifuged at 3500 rpm (1900 G) for 10 minutes and then plasma was collected and stored at −80°C until determination of plasma concentrations by the analytical laboratory.

ARI, DARI and OLA plasma concentrations were quantified by a high-performance liquid chromatography tandem mass spectrometry (HPLC-MS/MS) method developed and validated in our laboratory (Koller et al., 2019).

The calculation of pharmacokinetic parameters was explained in our previous study (Koller, Saiz-Rodríguez, et al., 2020).

### Pharmacodynamic Analysis

BP was measured in supine position with an automatic monitor at screening, predose, 16 times during treatment and 8 times after the last drug administration. Likewise, the 12-lead ECG was obtained at the same time points. QTc and HR were automatically calculated by the ECG device. Bazett correction formula was used to correct the QT interval (Bazett, 1997). According to the International Council for Harmonisation E14 clinical guidance (International Council on Harmonisation, 2005), a QTc interval greater than 450 milliseconds or a change from baseline greater than 30 milliseconds were considered as QTc interval prolongation.

### Safety and Tolerability Assessments

Safety and tolerability of ARI was assessed by clinical evaluation of adverse events (AEs) and other parameters including vital signs, physical examinations, and 12-lead ECGs. During the development of the study, volunteers were asked if they had experienced any AE. Moreover, the Ramsay sedation scale (Ramsay et al., 1974) was evaluated 4 times/day and on each safety visit, while the UKU side effect rating scale (Lingjærde et al., 1987) was evaluated on days 2, 4, 6, 9 and 15. According to the algorithm of the Spanish Pharmacovigilance System (Aguirre and García, 2016), the causality of AEs were classified as definite, probable, possible, unlikely or unrelated. Only those AEs that were definite, probable, or possible were considered as ADRs for statistical analysis. Intensity (mild, moderate, and severe), time sequence and outcome of AEs were also registered.

ADRs were classified using system organ class allocation as general (asthenia, fatigue, tiredness and gait alterations), cardiovascular (palpitations), gastrointestinal (constipation, nausea, vomiting, hyposalivation, hypersalivation, dry mouth and diarrhea), nervous system (akathisia, headache, difficulties with concentration, dizziness, paraesthesia, presyncope, syncope, tremor, somnolence and restless legs), psychiatric (restlessness, insomnia, anxiety, abnormal orgasm and nightmares), respiratory (epistaxis, hiccups, cough and sore throat), endocrine (galactorrhea), metabolic (lack of appetite, increased appetite and hyporexia), reproductive (dysmenorrhea, mastalgia and menstrual irregularity), skin (hair loss, pruritus, rash and sweating), musculoskeletal (shoulder pain, knee pain, neck pain, upper limb weakness, lumbalgia, cramps, back pain and leg pain), infections (cold), eye (photophobia) and investigations (increased liver enzymes) (Brown et al., 1999).

### Genotyping

DNA was extracted from 1 mL of peripheral blood samples using a MagNA Pure LC DNA Isolation Kit in an automatic DNA extractor (MagNa Pure® System, Roche Applied Science, Indianapolis, Indiana). Thenceforth, it was quantified spectrophotometrically in a NanoDrop® ND-1000 Spectrophotometer (Nanodrop Technologies, Wilmington, Delaware, USA) and the purity of the samples was measured by the A_260/280_ absorbance ratio.

Samples were genotyped with TaqMan assays using the OpenArray platform on a QuantStudio 12K Flex instrument (Thermo Fisher Scientific, Waltham, Massachusetts, USA). The genotyping array included 120 SNPs, whereof the following 46 were analysed in 14 genes based on their importance in the metabolism and mechanism of action of ARI and OLA: *CYP1A2* *1C (rs2069514), *1F (rs762551), *1B 5347T>C (rs2470890), *CYP2D6* *3 (rs35742686), *4 (rs3892097), *6 (rs5030655), *7 (rs5030867), *8 (rs5030865), *9 (rs5030656), *10 (rs1065852), *14 (rs5030865), *17 (rs28371706), *41 (rs28371725), *CYP3A4* *22 (rs35599367), rs55785340, rs4646438, *CYP3A5* *3 (rs776746), *6 (rs10264272), *ABCB1* C3435T (rs1045642), G2677 T/A (rs2032582), C1236T (rs1128503), rs3842, 1000-44G>T (rs10276036), 2895+3559C>T (rs7787082), 330-3208C>T (rs4728709), 2481+788T>C (rs10248420), 2686-3393T>G (rs10280101), 2320-695G>A (rs12720067), 2482-707A>G (rs11983225), 2212-372A>G (rs4148737), *ADRA2A* rs1800544, *BDNF* Val66Met (rs6265), *COMT* rs4680, rs13306278, *DRD2* TaqIA (rs1800497), 957C>T (rs6277), -141 Ins/Del (rs1799732), *DRD3* Ser9Gly (rs6280), *HTR2A* T102C (rs6313), C1354T (rs6314), rs7997012, *HTR2C* -759C/T (rs3813929), -697G/C (rs518147), rs1414334, *OPRM1* rs1799971 and *UGT1A1* rs887829.

Genotyping results were analysed within both the QuantStudio™ 12K Flex and Thermo Fisher Cloud softwares (Thermo Fisher Scientific, Waltham, Massachusetts, USA). Finally, a matrix of genotypic calls was exported for each polymorphism.

*CYP2D6* *29 (rs16947) polymorphism was genotyped in a 96-well thermal block installed in the same QuantStudio 12K flex instrument using individual TaqMan® probes. Additionally, *CYP3A4* *20 (rs67666821) polymorphism was genotyped by the KASPar SNP Genotyping System (LGC Genomics, Herts, UK). The ABI PRISM 7900HT Sequence Detection System (Thermo Fisher Scientific, Waltham, Massachusetts, USA) was used for fluorescence detection and allele assignment (Apellániz-Ruiz et al., 2015).

Copy number variations (CNVs) in the *CYP2D6* gene were determined with the TaqMan® Copy Number Assay (Assay ID: Hs00010001_cn; Thermo Fisher Scientific, Waltham, Massachusetts, USA) which detects a specific sequence on exon 9. Samples were run in a 96-well thermal block installed in the same QuantStudio 12K flex instrument

### Statistical analyses

Statistical analyses were performed with SPSS 24.0 software (SPSS Inc., Chicago, Illinois, United States). P values lower than 0.05 were considered statistically significant. The Hardy-Weinberg equilibrium was estimated for all genetic variants. Deviations from the equilibrium were detected by comparing observed and expected frequencies using a Fisher exact test based on the De Finetti program (available at http://ihg.gsf.de/cgi-bin/hw/hwa1.pl). Changes in BP, HR and QTc were analysed by repeated measures ANOVA. Pharmacokinetic parameters and polymorphisms were analysed as covariates. Estimate of effect size (partial eta squared, η_p_^2^), i.e. the proportion of the total variance that is attributed to an effect, is reported for each ANOVA test to avoid type II errors. Chi-square test was used to compare incidence of adverse effects between different genotypes. Bonferroni post-hoc test for multiple comparisons was applied for each analysis.

*CYP2D6* *3, *4, *5, *6, *7, *8, *9, *10, *14, *17, *29 and *41 were classified in phenotypes based on the functionality of the alleles (Caudle et al., 2020; Gaedigk et al., 2008). *CYP3A4* *2, *20, *22, and *CYP3A5* *3 and *6 genotypes were merged into a CYP3A phenotype (Sanchez Spitman et al., 2017). *CYP1A2* *1C, *1F and *1B variants were also merged into a phenotype (Saiz-Rodríguez, Ochoa, Belmonte, et al., 2019). *ABCB1* variants were merged into haplotypes: 0-8 mutant alleles were assigned to group 1, 9-12 mutant alleles were assigned to group 2 and 13-17 mutant alleles were assigned to group 3. Another *ABCB1* haplotype was assembled by only considering C3435T, G2677T/A and C1236T polymorphisms due to greater impact on the transporter’s activity or expression levels (Vivona et al., 2014). Zero or one mutant allele carriers were assigned to group 1, carriers of 2 or 3 mutant alleles were assigned to group 2 and bearing 4, 5 or 6 mutant alleles were assigned to group 3. *COMT* rs13306278 and rs4680 polymorphisms were merged into a haplotype: carrying no mutant allele was assigned as wild-type, carrying one mutant allele was considered as heterozygous while carrying more than one mutant allele was considered as mutant.

## Results

### Demographic characteristics and genotype frequencies

Demographic data are shown in *Table* 1. Ten subjects were Caucasian and 14 were Latin. Age was similar between males and females. Males had greater weight and height than females (*p* < 0.001), however, BMI values did not differ significantly (*p* = 0.798).

**Table 1.** Demographic characteristics.

|  | N (%) | Age (y) | Weight (kg) | Height (m) | BMI (kg/m<sup>2</sup>) |
| --- | --- | --- | --- | --- | --- |
| **All** | 24 (100) | 31.5 ± 11.6 | 71.4 ± 12.2 | 1.68 ± 0.11 | 25.3 ± 2.6 |
| **Males** | 12 (50) | 28.5 ± 7.4 | 78.4 ± 12.2 | 1.76 ± 0.09 | 25.4 ± 2.8 |
| **Females** | 12 (50) | 34.6 ± 14.3 | 64.3 ± 7.4* | 1.60 ± 0.07* | 25.1 ± 2.5 |

Values are shown as mean ± SD unless otherwise indicated. \*p < 0.05 versus males.

Genotype frequencies of the analysed genes are shown in *Table* S1.

### Pharmacokinetics

Mean and standard deviation (SD) of ARI, DARI and OLA pharmacokinetic parameters are shown in *Table* 2. Females had lower DARI/ARI ratio than males (*p* = 0.046). The remaining pharmacokinetic parameters were not statistically different between sexes.

**Table 2.**
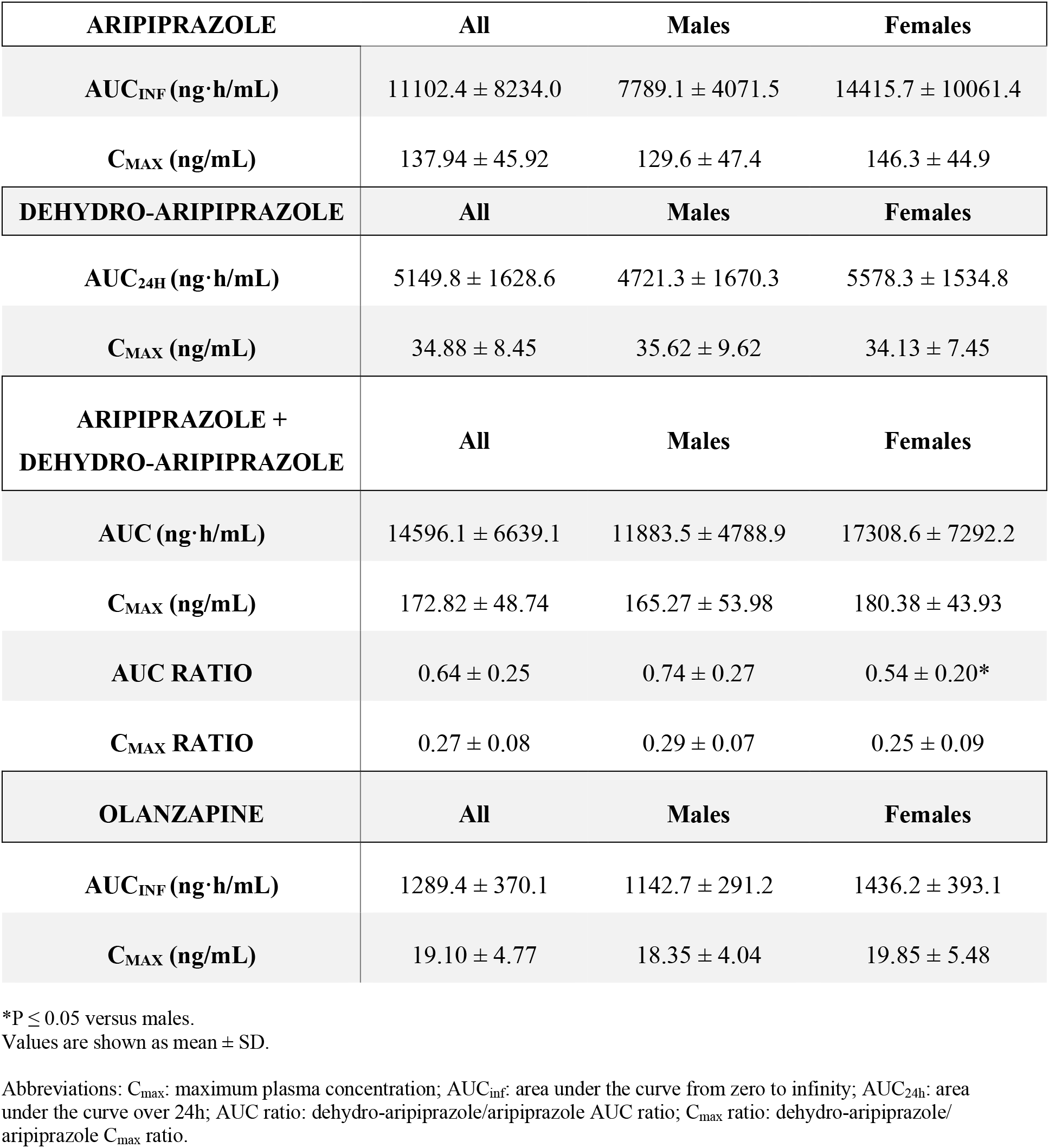
Pharmacokinetic parameters of aripiprazole, dehydro-aripiprazole and olanzapine after 5 days administration of aripiprazole 10 mg/day or olanzapine 5 mg/day.

| **ARIPIPRAZOLE** | **All** | **Males** | **Females** |
| --- | --- | --- | --- |
| **AUC<sub>INF</sub> (ng·h/mL)** | 11102.4 ± 8234.0 | 7789.1 ± 4071.5 | 14415.7 ± 10061.4 |
| **C<sub>MAX</sub> (ng/mL)** | 137.94 ± 45.92 | 129.6 ± 47.4 | 146.3 ± 44.9 |
| **DEHYDRO-ARIPIPRAZOLE** | **All** | **Males** | **Females** |
| **AUC<sub>24h</sub> (ng·h/mL)** | 5149.8 ± 1628.6 | 4721.3 ± 1670.3 | 5578.3 ± 1534.8 |
| **C<sub>MAX</sub> (ng/mL)** | 34.88 ± 8.45 | 35.62 ± 9.62 | 34.13 ± 7.45 |
| **ARIPIPRAZOLE +**<br>**DEHYDRO-ARIPIPRAZOLE** | **All** | **Males** | **Females** |
| **AUC (ng·h/mL)** | 14596.1 ± 6639.1 | 11883.5 ± 4788.9 | 17308.6 ± 7292.2 |
| **C<sub>MAX</sub> (ng/mL)** | 172.82 ± 48.74 | 165.27 ± 53.98 | 180.38 ± 43.93 |
| **AUC RATIO** | 0.64 ± 0.25 | 0.74 ± 0.27 | 0.54 ± 0.20* |
| **C<sub>MAX</sub> RATIO** | 0.27 ± 0.08 | 0.29 ± 0.07 | 0.25 ± 0.09 |
| **OLANZAPINE** | **All** | **Males** | **Females** |
| **AUC<sub>INF</sub> (ng·h/mL)** | 1289.4 ± 370.1 | 1142.7 ± 291.2 | 1436.2 ± 393.1 |
| **C<sub>MAX</sub> (ng/mL)** | 19.10 ± 4.77 | 18.35 ± 4.04 | 19.85 ± 5.48 |

\*P ≤ 0.05 versus males.
Values are shown as mean ± SD.
Abbreviations: C<sub>max</sub>: maximum plasma concentration; AUC<sub>inf</sub>: area under the curve from zero to infinity; AUC<sub>24h</sub>: area under the curve over 24h; AUC ratio: dehydro-aripiprazole/aripiprazole AUC ratio; C<sub>max</sub> ratio: dehydro-aripiprazole/aripiprazole C<sub>max</sub> ratio.

### Pharmacodynamics

#### Aripiprazole

ARI had a DBP lowering effect on the first day of treatment (5 mmHg, *p* = 0.004, η_p_^2^ = 0.309) and a QTc lowering effect on days 3 and 5 (20 ms, *p* < 0.001, η_p_^2^ = 0.512; 15 ms, *p* = 0.028, η_p_^2^ = 0.193, respectively) (*Table* S2). None of the volunteers had a QTc value higher than 450 ms or showed more than a 30 ms change. HR incremented from predose to 5 h after drug administration on the 4^th^ day in males, while it did not change in females (*p* = 0.034, η_p_^2^ = 0.197). DBP on the first day decreased more in *HTR2A* rs6313 C allele carriers and in *ADRA2A* rs1800544 C/C subjects compared to T/T and C/G subjects (7 vs +3 mmHg, *p* = 0.006, η_p_^2^ = 0.296; 7 vs 0 mmHg, *p* = 0.020, η_p_^2^ = 0.224, respectively).

#### Olanzapine

OLA had a SBP, DBP, HR and QTc lowering effect on the first day 5 h after drug administration (16 mmHg*, p* < 0.001, η_p_^2^ = 0.678; 8 mmHg, *p* < 0.001, η_p_^2^ = 0.537; 10 bpm*, p* < 0.001, η_p_^2^ = 0.129; 10 ms, *p* = 0.002, η_p_^2^ = 0.359, respectively) (*Table* S3). None of the volunteers had a QTc value higher than 450 ms or showed more than a 30 ms change. HR increased from predose to 5 h after drug administration on the 4^th^ day in males, while it did not change in females (*p* = 0.002, η_p_^2^ = 0.350). On the 4^th^ day of drug administration, QTc values increased in males and the opposite effect was detected in females (*p* = 0.018, η_p_^2^ = 0.227). SBP on the first day decreased more in *DRD3* rs6280 Ser/Ser and Ser/Gly and in *ADRA2A* rs1800544 C/C subjects compared to volunteers with Gly/Gly and C/G genotypes (21 and 14 vs 6 mmHg, *p* = 0.025, η_p_^2^ = 0.209; 18 vs 8 mmHg, *p* = 0.048, η_p_^2^ = 0.167, respectively). Moreover, DBP on the first day diminished more in *COMT* wild-type subjects compared to those with heterozygous and mutant phenotype (16 vs 8 and 5 mmHg, *p* = 0.022, η_p_^2^ = 0.218). Additionally, HR on the first day decreased more in *DRD3* rs6280 Ser/Ser and Ser/Gly subjects compared to those with Gly/Gly genotype (16 and 15 vs 4 mmHg, respectively, *p* = 0.013, η_p_^2^ = 0.249).

The effects of ARI and OLA on BP, HR and QTc are compared in *Figure* 1. Changes in SBP, DBP and QTc were not statistically significant between ARI and OLA (*p* = 0.110, *p* = 0.145 and *p* = 0.236, respectively). However, although it did not reach the statistically significant level, OLA lowered both SBP and DBP to a greater extent compared to ARI. After the 4^th^ day, tolerance was developed for the hypotensive effect of OLA. Additionally, HR was significantly lower during OLA treatment compared to ARI (*p* < 0.0001, η_p_^2^ = 0.236).

**Figure 1.**
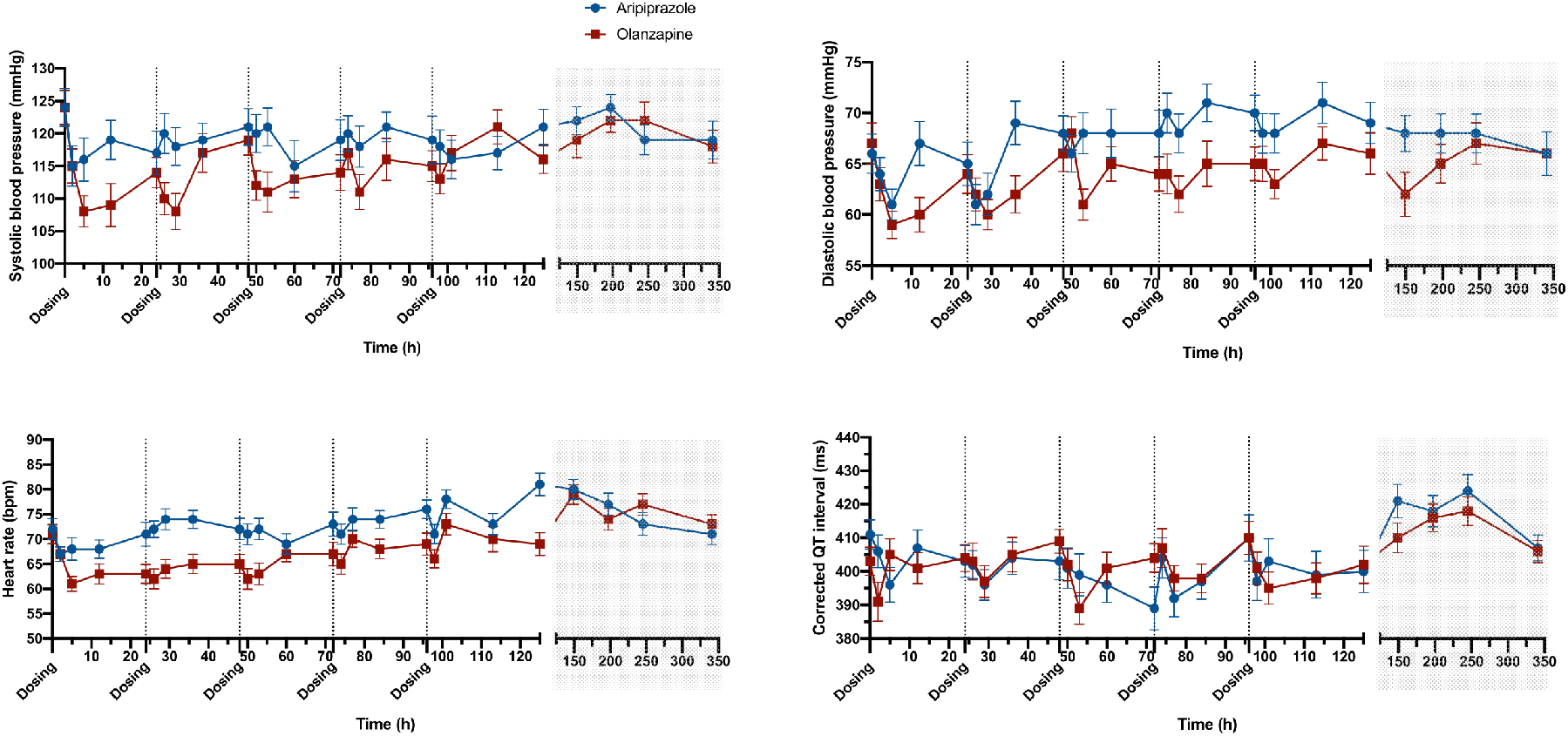
Effects of aripiprazole and olanzapine on blood pressure, heart rate and corrected QT interval. The shaded section shows the values after the end of treatment. The results are shown in mean ± SD.

### Adverse drug reactions

During the study, no serious or life-threatening AEs were documented. All volunteers experienced at least one ADR.

### Aripiprazole

The most frequent ADRs were somnolence (79%), headache (54%), insomnia (33%), dizziness (21%), restlessness (21%), palpitations (21%), akathisia (17%) and nausea (17%) (*Table* 3). The number of ADRs was similar between males and females (3.9 ± 2.3 and 4.1 ± 2.0 ADR/ subject, respectively). Palpitations were only registered in females (5 volunteers, *p* = 0.012 compared to males). The incidence of akathisia was higher in *DRD3* rs6280 Ser/Ser homozygotes compared to Gly allele carriers (22.2% vs 0%, respectively, *p* = 0.003). Moreover, only *DRD2* rs1799732 G/-subjects experienced asthenia compared to G/G homozygotes (33.3% vs 0%, respectively, *p* = 0.011). The incidence of headache was significantly higher in *HTR2C* rs3813929 T carriers than in C/C homozygotes (66.7% vs 28.6%, respectively, *p* = 0.030). Additionally, an association was found between CYP1A2 NM/RM phenotype and the incidence of insomnia (41.2% vs 0% in UM, *p* = 0.044). Finally, somnolence was detected more frequently in *HTR2A* rs6314 C/C and *OPRM1* rs1799971 A/A subjects compared to C/T subjects and G carriers, respectively (86.4% vs 0%, *p* = 0.004 and 89.5% vs 40%, *p* = 0.029, respectively).

**Table 3.** Adverse drug reactions to aripiprazole and olanzapine.

| Adverse event | Aripiprazole<br>N (%) | Olanzapine<br>N (%) |
| --- | --- | --- |
| **Nervous system disorders** |  |  |
| Akathisia | 4 (17) | - |
| Dizziness | 5 (21) | 7 (29) |
| Headache | 13 (54) | 3 (13) |
| Restless legs | 1 (4) | - |
| Somnolence | 19 (79) | 24 (100) |
| Pre-syncope | - | 1 (4) |
| Syncope | 1 (4) | 1 (4) |
| Left hand tremor | - | 1 (4) |
| **Psychiatric disorders** |  |  |
| Anxiety | 2 (8) | - |
| Early awakening | 1 (4) | - |
| Restlessness | 5 (21) | 1 (4) |
| Insomnia | 8 (33) | 2 (8) |
| **General disorders** |  |  |
| Asthenia | 2 (8) | 3 (13) |
| Fatigue | 2 (8) | - |
| Gait alterations | 1 (4) | - |
| Tiredness | 1 (4) | - |
| Weakness | 1 (4) | 1 (4) |
| **Gastrointestinal disorders** |  |  |
| Constipation | 3 (13) | 3 (13) |
| Dry mouth | - | 3 (13) |
| Gastric discomfort | 1 (4) | - |
| Hypersalivation | - | 1 (4) |
| Hyposalivation | 1 (4) | - |
| Nausea | 4 (17) | 3 (13) |
| Vomiting | 2 (8) | - |
| **Skin and subcutaneous tissue disorders** |  |  |
| Facial rash | 1 (4) | 1 (4) |
| Hair loss | 1 (4) | - |
| Rash | - | 1 (4) |
| Sweating | 3 (13) | - |
| Pruritus | - | 1 (4) |
| **Respiratory, thoracic and mediastinal disorders** |  |  |
| Hiccups | 3 (13) | - |
| **Metabolism and nutrition disorders** |  |  |
| Hyporexia | 1 (4) | - |
| Increased appetite | - | 1 (4) |
| **Investigations** |  |  |
| Increased liver enzymes | 2 (8) | 1 (4) |
| **Cardiac disorders** |  |  |
| Palpitations | 5 (21) | 2 (8) |
| **Musculoskeletal and connective tissue disorders** |  |  |
| Upper limb weakness | 1 (4) | - |
| Left shoulder pain | - | 1 (4) |
| **Eye disorders** |  |  |
| Photophobia | - | 1 (4) |

### Olanzapine

The most frequent ADRs were somnolence (100%), dizziness (29%), asthenia (13%), constipation (13%), dry mouth (13%), headache (13%) and nausea (13%) (*Table* 3). The number of ADRs was similar between males and females (1.7 ± 1.8 and 1.7 ± 1.1 ADR/ subject, respectively). Constipation was detected more frequently in *HTR2A* rs6314 T allele carriers, *HTR2A* rs7997012 A allele carriers and *UGT1A1* rs887829 T/T subjects compared to C/C homozygotes, G/G homozygotes and C allele carriers (100.0% vs 4.5%, *p* < 0.001; 15.4% vs 9.0%, *p* = 0.026 and 60.0% vs 0%, *p* = 0.001, respectively). Moreover, only *DRD3* rs6280 Ser/Ser subjects experienced dry mouth compared to Gly allele carriers (50.0% vs 0%, respectively, *p* = 0.006). The incidence of insomnia was higher in *HTR2C* rs1414334 G/G homozygotes compared to C allele carriers (100.0% vs 4.2%, respectively, *p* = 0.003). In addition, nausea was only detected in *HTR2C* rs518147 C/T heterozygotes and not in C/C and T/T homozygotes (33.3% vs 0%, respectively, *p* = 0.038). Finally, palpitations were only reported in CYP1A2 UM and *HTR2A* rs7997012 A allele carriers and not in NM/RMs and G/G homozygotes (28.6% vs 0%, *p* = 0.021 and 15.4% vs 0%, *p* = 0.002, respectively).

ADRs to ARI and OLA classified by system organ class allocation are shown in *Figure* 2. The number of registered ADRs was significantly higher after ARI administration (91 vs 60, *p* < 0.035). Likewise, more psychiatric and cardiac ADRs were detected during ARI treatment compared to OLA (16 vs 3 and 7 vs 2, respectively, *p* < 0.001).

**Figure 2.**
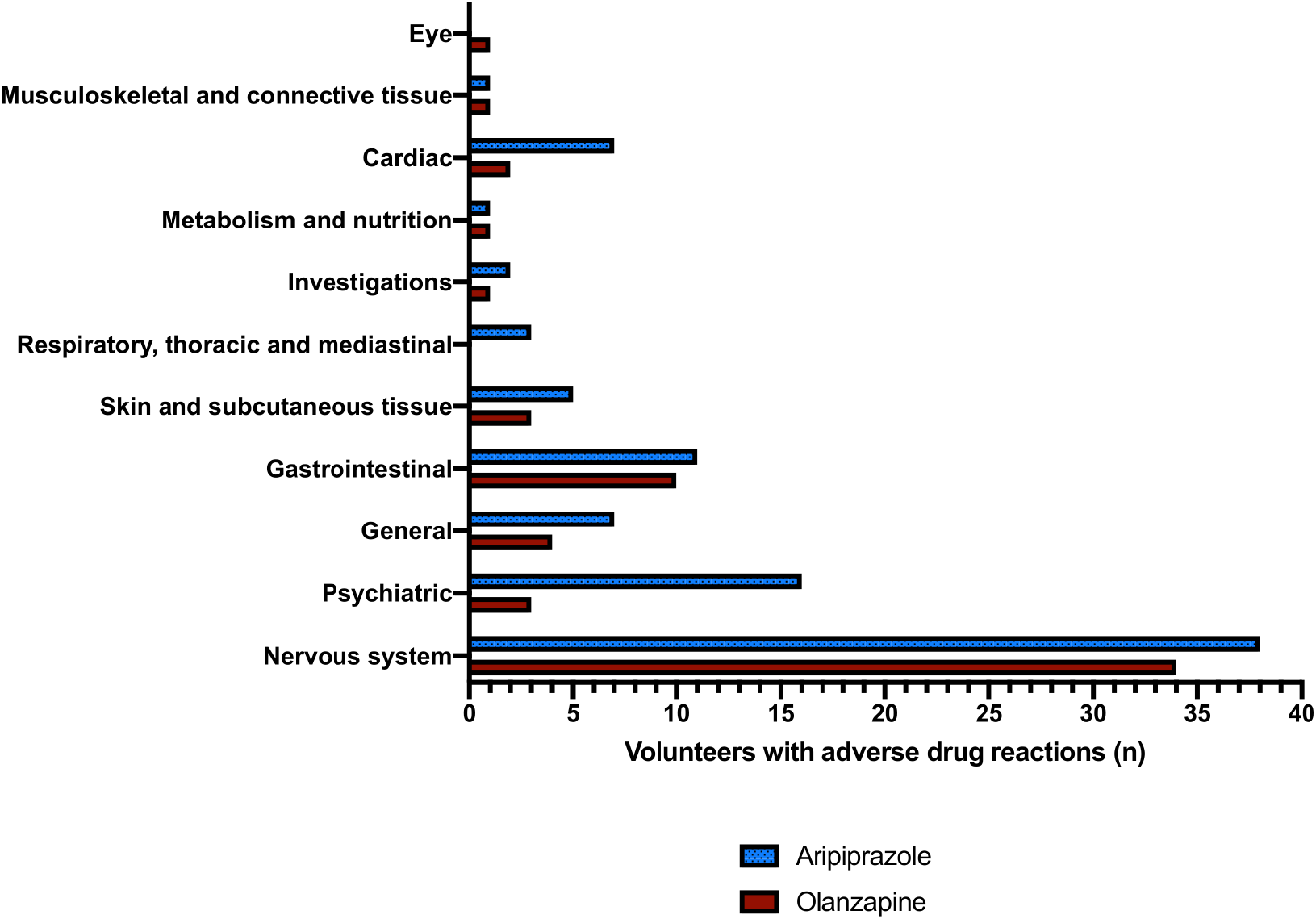
Adverse drug reactions to aripiprazole and olanzapine classified in groups.

## Discussion

### Pharmacodynamics

#### Effects on the blood pressure and heart rate

ARI caused both hyper- and hypotension in previous studies. Hypertension was reported in case reports. The elevated blood pressure dropped immediately after ARI withdrawal (Yasui-Furukori and Akira Fujii, 2013). In contrast, in other studies, dose-related hypotension was also reported: when ARI was reduced to 5 mg/day (Wang, 2013) or discontinued (Torgovnick et al., 2008), the blood pressure returned to normal range. On the contrary, OLA has little effects on the cardiovascular system, if any (Khasawneh and Shankar, 2014). Nevertheless, it can cause hypotension (Lee et al., 2003). Bradycardia was also reported previously during OLA treatment (Lee et al., 2003; Markowitz et al., 2002).

In our study, ARI decreased SBP (without reaching significance) and DBP on the first day of treatment, but this effect was not repeated on the rest of the days. The BP lowering mechanism may be due to blocking the α1-adrenergic receptors. Moreover, its 5-HT2A antagonism could induce vasodilation and its 5-HT1A agonism could produce hypotension and bradycardia (Lin et al., 2017). Regarding OLA, SBP, DBP and HR significantly decreased after the first dose. These changes could be explained by its α1-adrenergic antagonism (Beasley et al., 1996). These effects were only significant on the first day of treatment and progressively diminished on the following days (*Figure* 1) as tolerance was developed. It was reported previously that only the first dose of OLA caused hypotension and bradycardia (Bever and Perry, 1998). Our study is the first to report this association with ARI.

SBP and DBP decreased more in *ADRA2A* rs1800544 C/C subjects compared to C/G subjects during OLA and ARI treatment, respectively. α2A-adrenergic receptors also have important roles in sympathetic cardiovascular regulation. Mice that do not express *ADRA2A* had increased BP and HR (Kurnik et al., 2011). Consequently, rs1800544 mutant allele carriers should have increased BP compared to wild-type homozygotes.

DBP decreased more in *HTR2A* rs6313 C allele carriers compared to T/T subjects during ARI treatment. Carriers of the wild-type allele of *HTR2A* rs6313 could have induced vasodilatation, and therefore a decrease in blood pressure (Lin et al., 2017).

During OLA treatment, SBP and HR decreased more in *DRD3* rs6280 Ser/Ser and Ser/Gly subjects compared to those with Gly/Gly genotype. Dopamine causes cardiac stimulation and therefore vasoconstriction and increase in BP. OLA blocks dopamine receptors, therefore less dopamine binds to them what can result in decreased BP and HR (Bangash et al., 2012). D3 dopamine receptor blockage depends on the genotype, what explains that carriers of the mutant genotype may have a less efficient interaction between the drug and the receptor, causing smaller variations in BP and HR.

In previous studies, *COMT* rs4680 A (Val, wild-type) allele carriers had lower SBP and DBP throughout the study (Ge et al., 2015; Htun et al., 2011). Our study confirmed these findings: subjects with the COMT wild-type phenotype (including COMT rs4680) had significantly higher DBP decrease after OLA administration. However, another study found the opposite: the wild-type allele was associated with SBP elevation (Hagen et al., 2007). Thus, there is no clear consensus about the role of *COMT* polymorphisms in its BP-lowering capacity. Nevertheless, this association could be due to its role in modulating dopamine function (Tunbridge, 2010).

Habitually, females show higher HR and QTc than males (Li et al., 2013; Lutfi and Sukkar, 2011). In our previous study both were higher in females after a single dose of ARI (Belmonte et al., 2016). In our current study we found the contrary with ARI and OLA, however, as this difference could only be seen on the 4^th^ day of drug administration, it may be considered an artefact.

#### Effects on corrected QT interval

The mean QTc interval normally decreases with ARI and the QTc prolongation risk is lower compared to other atypical antipsychotics (Polcwiartek et al., 2015). The QTc interval was overall decreased in case reports and clinical trials including schizophrenic patients and healthy volunteers, however, QTc prolongation events were also discovered (Polcwiartek et al., 2015). Our study confirms that ARI induces QTc decrease what started on the second day of drug administration and was maintained forth until the last day. Therefore, based on our results, ARI does not seem to cause QTc prolongation and consequently *Torsades de Pointes* and sudden cardiac death. OLA does not induce QTc prolongation (Czekalla et al., 2001). Based on the authors knowledge, the current study is the first to report QTc decrease. However, it was produced only on the first day of drug administration alike BP and HR. To date, the clinical significance of its QTc-shortening is uncertain. Nevertheless, it may induce proarrhythmia (Shah, 2010).

### Adverse drug reactions

#### Most common adverse drug reactions to ARI

According to the drug label, the most common ADRs to ARI in schizophrenic patients are akathisia, extrapyramidal symptoms, somnolence and tremor (Ribeiro et al., 2018). Somnolence and akathisia were among the most frequent ADRs that we observed. The most common ADRs in clinical trials in healthy volunteers and psychotic patients were nausea, vomiting, constipation, headache, dizziness, akathisia, anxiety, insomnia, and restlessness (Ribeiro et al., 2018). All these ADRs were detected in our clinical trial in 17, 8, 13, 54, 21, 17, 8, 33 and 21% of the volunteers, respectively. There is no evidence on sex differences in the prevalence of ADRs (Montero et al., 2008). Our study confirms this hypothesis. In general, the most common ADRs to ARI were nervous system and psychiatric conditions.

#### Most common adverse drug reactions to OLA

According to the drug label, the most common ADRs to OLA in schizophrenic patients are constipation, weight gain, dizziness, personality disorder, akathisia, postural hypotension, sedation, headache, increased appetite, fatigue, dry mouth and abdominal pain (R. R. Conley and Meltzer, 2000). Constipation, dizziness, headache and dry mouth were among the most frequent ADRs that we observed in 13, 29, 13 and 13% of the volunteers, respectively. ADRs that we could not detect in our clinical trial were personality disorder, fatigue, sedation and abdominal pain. The absence of these findings could be due to the short treatment; the majority of these ADRs usually appear after at least 6 weeks of treatment (R. R. Conley and Meltzer, 2000). A mild weight gain was observed during our study which was reported in our previous article (Koller, Almenara, et al., 2020). There are possible sex differences in the prevalence of ADRs to OLA (Seeman, 2009). However, we did not see a difference between the two sexes in our study. The lack of association could be due to the short-time treatment and the low sample size. The most common ADRs to OLA were metabolic disorders.

#### Cardiac disorders

Palpitations are considered infrequent ADRs to ARI (Ribeiro et al., 2018). However, 5 female volunteers and no males experienced it in our study. Palpitations may also occur during OLA treatment, however, they are not considered as frequent ADRs (Chaves et al., 2013). Our study confirms this finding; only 2 volunteers were registered with palpitations. Interestingly, both were females, similar to ARI. We are the first to report these findings. Palpitations can be associated with arrhythmias. The proarrhythmic effect, drug-induced *Torsade de Pointes* occurs more frequently in females than in males (Wolbrette et al., 2002) and was observed with ARI (Polcwiartek et al., 2015), but not with OLA (Glassman and Bigger, 2001). Therefore, as in the current study, palpitations, a proarrhythmic sign, may occur more frequently in females.

In addition, during OLA treatment palpitations were only reported in CYP1A2 UMs and *HTR2A* rs7997012 A allele carriers. OLA is metabolized by CYP1A2 (Callaghan et al., 1999). UMs may reach high concentrations of OLA metabolites rapidly, which could be related to the development of palpitations. Additionally, palpitations were only reported in *HTR2A* rs7997012 mutant (A) allele carriers. Serotonin can induce the development of palpitations (Trindade et al., 1998). Therefore, A allele carriers may have reduced 5-HTR2A blockage and consequently higher serotonin levels.

#### Nervous system disorders

Akathisia is commonly associated with first generation antipsychotics. It would be expected that ARI had a low incidence of akathisia being an antagonist at 5-HT2A receptors (Thomas et al., 2015). However, ARI seems to increase the risk of akathisia. Therefore, its pathophysiology seems complex, involving several neurotransmitters including dopamine, acetylcholine, γ-aminobutyric acid, norepinephrine, serotonin and neuropeptides (Iqbal et al., 2007). Consequently, *DRD3* mutant (Gly) allele carriers could be more protected from developing akathisia. The fact that *DRD2* rs1799732 G/- subjects experienced asthenia but not G/G homozygotes strengthens the dopamine theory: polymorphisms in its receptors seem to have a role in developing nervous system ADRs. On the contrary, OLA does not cause akathisia (Thomas et al., 2015) what was confirmed in our study. It appears that its sedating properties could be responsible for attenuating the effects of akathisia (Thomas et al., 2015).

5-HT receptors are related to the development of somnolence and headache during antipsychotic treatment, but the molecular background is unknown to date (Fang et al., 2016; LaPorta, 2007). ARI relates to a lower risk of somnolence and headache compared to other atypical antipsychotics (Fang et al., 2016), however, they are still among the most common ADRs (Ribeiro et al., 2018). Somnolence was detected more frequently in *HTR2A* rs6314 wild-type subjects while headache was observed more in mutant allele carriers of *HTR2C* rs3813929. Our study is the first to report these findings. It strengthens the hypothesis that 5-HT receptor variability can lead to the development of these ADRs.

In our previous study, *OPRM1* rs1799971 mutant (G) allele carriers were associated with the increased likelihood of somnolence to fentanyl in healthy volunteers (Saiz-Rodríguez, Ochoa, Herrador, et al., 2019). In the current study we observed the contrary: wild-type (A/A) homozygous subjects developed somnolence more frequently during ARI treatment. The G variant is reported as a protective allele against ADRs (Matic et al., 2017). However, its role in developing ADRs to ARI is currently unknown. Opioid receptor activation inhibits GABAergic interneurons in order to increase dopamine release (Hirasawa-Fujita et al., 2017). Therefore, higher dopamine concentration in *OPRM1* rs1799971 wild-type homozygotes may increase the risk of somnolence.

In addition, CYP1A2 NM/RM subjects showed a higher prevalence of insomnia during ARI treatment than UM subjects. Based on current knowledge, ARI is not metabolized by CYP1A2. However, in our population, CYP1A2 UMs showed lower ARI and DARI disposition compared to the other phenotypes (Koller, Saiz-Rodríguez, et al., 2020). Therefore, NM/RM subjects were under prolonged ARI exposure what could cause the development of insomnia. Regarding OLA, insomnia is not among the most common ADRs to OLA (ZYPREXA (olanzapine), FDA., 1996). However, 2 volunteers experienced it during our study, both carrying *HTR2C* rs1414334 G/G genotype. The lack of serotonin can cause insomnia (Murray et al., 2015). *HTR2C* rs1414334 mutant (G/G) homozygotes may have less 5-HTR2C receptor blocking effect and consequently higher risk to experience insomnia.

#### Gastrointestinal disorders

Serotonin and acetylcholine activate the colonic smooth muscles inducing their contraction. OLA, being a 5-HT antagonist, inhibits their contraction and consequently causes constipation (Zhang et al., 2016). Carriers of *HTR2A* rs6314 and rs7997012 mutant alleles (T and A, respectively) displayed a higher incidence of constipation, therefore, in these subjects, OLA possibly has higher binding affinity to 5-HT2A receptors. In our previous study, the prevalence of fatigue in *UGT1A1* rs887829 T/T subjects was significantly higher compared to wild-type (C) allele carriers (Cabaleiro et al., 2013). In the current study, we found the same association, but with constipation. This enzyme may be responsible for the phase II metabolism of OLA (Koller, Saiz-Rodríguez, et al., 2020). These associations were not found with ARI what we expected as it is not metabolized by the UGT enzyme family. However, similar to OLA, it is a 5-HT2A antagonist. Nevertheless, this mechanism should be more complex; dopamine or other neurotransmitters could have a role in the development of constipation.

Normally, a balance is maintained between acetylcholine and dopamine. When this balance is disturbed, acetylcholine levels increase, while dopamine levels decrease (Aosaki et al., 2010). Dry mouth is an anticholinergic side effect of OLA, but not to ARI given its special mechanism of action (Bhana et al., 2001). *DRD3* rs6280 wild-type (Ser/Ser) subjects may have higher dopamine level due to the low amount of acetylcholine, therefore experiencing dry mouth after OLA.

Nausea during OLA treatment was only detected in *HTR2C* rs518147 C/T heterozygotes and not in C/C and T/T homozygotes what can be due to the low sample size. This association was not found with ARI.

## Study limitations

The main limitation of our study is the low sample size. Therefore, it is of importance to interpret these results with caution. Our results should be confirmed in further studies with healthy volunteers to increase the sample size as well as in schizophrenic patients to demonstrate their clinical utility. Notwithstanding, our conditions were perfectly controlled and we administered both drugs to each volunteer, what can reduce the influence of other factors, such as comorbility, smoking and nutrition. Additionally, the duration of this study was short and several ADRs to ARI and OLA could appear later than 5 days. However, the Ethics Committee would rarely authorize more than 5 days treatment with antipsychotics in healthy volunteers.

## Conclusions

OLA had more cardiovascular effects than ARI. However, BP, HR and QTc decreased significantly only on the first day of drug administration. Therefore, it seems that a rapid tolerance is developed to the drug. We suggest that *HTR2A, ADRA2A, DRD3* and *COMT* polymorphisms establish the interindividual variability of the cardiovascular effects of ARI and OLA. Contrastively, more ADRs were registered to ARI than to OLA, especially psychiatric and nervous system disorders. Additionally, we propose that *HTR2A, HTR2C, DRD2, DRD3, OPRM1, UGT1A1* and *CYP1A2* polymorphisms have a role in the development of ADRs to ARI and OLA. Consequently, some polymorphisms may explain the difference in the incidence of ADRs among subjects.

## Data Availability

The details about the study are available on the webpage of the Spanish Agency of Medicines and Health Products.

https://reec.aemps.es/reec/public/detail.html

## Acknowledgements

The authors are grateful to the volunteers as well as the effort of the staff of the Clinical Trials Unit of our hospital, especially to Alejandro de Miguel-Cáceres for sample processing and Maria Hernández and Diana Campodónico for safety evaluations.

## Author Contributions

Wrote manuscript: Dora Koller; Designed research: Dora Koller, Francisco Abad-Santos, Miriam Saiz-Rodríguez, Dolores Ochoa; Performed clinical trial: Manuel Román, Gina Mejía, Francisco Abad-Santos, Daniel Romero-Palacián, Samuel Martín, Dolores Ochoa; Analysed data: Dora Koller, Susana Almenara, Gina Mejía, Francisco Abad-Santos; Determination of drug concentrations: Dora Koller, Pablo Zubiaur, Aneta Wojnicz; Pharmacogenetic assessments: Dora Koller, Pablo Zubiaur, Marcos Navares, Miriam Saiz-Rodríguez.

